# ASSESSMENT OF RELATIONSHIP BETWEEN CLINICAL MANIFESTATION OF CHIARI MALFORMATION TYPE I AND CEREBELLAR TONSILS HERNIATION MEASUREMENT WITHIN THE FORAMEN MAGNUM

**DOI:** 10.1101/2023.04.23.23288530

**Authors:** Shingiro Eric, Munyemana Paulin, Inyange Sylvie, Rudakemwa Emmanuel, Muneza Severien, Hakizimana David, Nkusi Agaba Emmy

**Affiliations:** University Of Rwanda College of Medicine and Health Sciences, Department of Surgery; Rwanda Military Hospital, Department of Neurosurgery; Kigali University Teaching Hospital, Department Of Neurosuregry; King Faisal Hospital, Kigali

**Keywords:** Chiari malformation type I, headache and myelopathy severity, cerebellar tonsils volume and T/F volume ratio within foramen magnum

## Abstract

Chiari malformation type I(CMI) is a common condition. It is a subject of controversy from diagnosis to the management (16). Classically the diagnosis is made on clinical basis and radiological measurement of cerebellar tonsils herniation of 5mm or more below the opisthion-basion line in mid-sagittal plane(Mc Rae line.) The aim of our study was to determine the relationship between clinical presentation of CMI and cerebellar tonsil herniation measured in three dimensions, cerebellar tonsils volume and the volume ratio (cerebellar tonsils volume/Foramen magnum volume) within foramen magnum. Can the volume of cerebellar tonsils herniation and the volume ratio(cerebellar tonsils volume/volume foramen magnum) reflect better the severity of patients with CMI? the study is the first in current literature eliciting the relationship between myelopathy severity and headache severity in CMI patients; cerebellar tonsils volume and T/F volume ratio (cerebellar tonsils volume /Foramen magnum volume)

**Methods:** We conducted an observational cross sectional analytical study. Patients with clinical and radiological confirmation of CMI evaluated on cranial cervical MRI were enrolled. Three dimension morphometric measures of cerebellar tonsils was made, the volume of cerebellar tonsils was calculated using ellipsoid volume formula. The transverse diameter of foramen magnum was measured and the volume of foramen magnum was calculated using sphere formula. We computed various non-parametric statistical tests and hypothesis testing to analyze variation of cerebellar tonsils uniplanar measurements, cerebellar tonsils volume, and T/F volume ratio (Cerebellar tonsils volume/foramen magnum volume), and to analyze correlation between these measurements with the severity of myelopathy using modified Japanese orthopedics association score(mJOA) and headache severity using pain numeric rating scale. We did all the calculations in python 3 using scipy. stats, Wilcoxon, Pearson, seaborn, and matplotlib.pyplot packages and pandas library

**Results:** Chiari malformation type I was more common in female with 61.5% and male patients with CMI was 38.5%. The majority of patients with CMI were in fourth and fifth decade. Occipital headache was the most presenting symptom followed by limb paraesthesia, vertigo, difficulty walking and bulbar symptoms. Scoliosis associated with CMI was found in 5% while syringomyelia associated with CMI was found in 8%. According to numeric pain scale; patients with CMI mostly presented with severe headache and moderate headache with 58.3% and 41.7 % respectively.

There is difference between right and left sagittal tonsils measurement; the left median sagittal measurement is 7.8 mm while the right median sagittal measurement is 8.8 mm with P-value <0.001

The coronal and sagittal cerebellar tonsils measurements are statistically different. The median difference and interquartile range(IQR) between coronal and sagittal measurements were 0.6(-0.4 1.8) and p-value <0.001 respectively

The finding showed a correlation between myelopathy severity and the volume of herniated cerebellar tonsils as well as correlation between myelopathy severity and T/F volume ratio (Cerebellar tonsils volume/Foramen magnum volume). There was no correlation between headache severity and sagittal measurement as we failed to reject hypothesis p=0.661 Spearman’s correlation coefficient: -0.045 In contrast there was a correlation between headache severity and cerebellar tonsils volume as well as T/F volume ratio with P-value 0.03 in our study.

**Conclusion:** Two dimensions radiological measurements in assessment of CMI is not reflecting the clinical severity of patients with CMI. Consideration of both clinical presentation and radiological measurement in assessment of severity of CMI is of great importance rather than only considering the cut off 5 mm descent of cerebellar tonsils herniation in midsaggital plan. Cerebellar tonsils volume and T/F volume ratio(cerebellar tonsils volume /foramen magnum volume) are the indicators of severity of myelopathy and headache severity as shown in our study.

## BACKGROUND

The description of CMI was initially based on pathological and anatomical anomaly related to the cerebellar tonsils (1). Based on anatomical definition; CMI is characterized by hindbrain deformity and elongation of the cerebellar tonsils resulting into their descent into the foramen magnum. The classic definition of CMI is radiological and is defined as herniation of the cerebellar tonsils below the foramen magnum of > 3 mm in children and> 5 mm in adults(2). There are questions about the most accurate radiological criteria to diagnose CMI which remain unanswered. Is CMI a sagittal disease? should we only rely on the cut off descent of cerebellar tonsils of 5mm below foramen magnum in sagittal plane in diagnosis of CMI ? can the descent of cerebellar tonsils in coronal plane reflect the severity of CMI? Can the volume of cerebellar tonsils herniation reflect better the severity of CMI ? Can T/F volume ratio (Cerebellar tonsil volume/Foramen magnum volume) reflect better the severity CMI patients.

## METHODS

This study was conducted in 3 teaching hospitals: King Faisal Hospital(KFH)-,University Teaching Hospital of Kigali (CHUK) and Rwanda Military Hospital(RMH). All of these hospitals are major tertiary referral hospitals having neurosurgeons treating CMI. Cerebellar tonsils measurements were taken from CIM Patients who underwent cranial cervical 1.5T MRI at King Faisal Hospital 1.5T during the study period. We conducted a prospective observational cross sectional analytical study. Patients with clinical and radiological confirmation of CMI were enrolled based on most common clinical findings in patients with CMI; clinical inclusion criteria was at least patients presenting with occipital headache or neck pain, dizziness, limb paraesthesia and increased deep tendon reflex 3+ on physical exam. Cerebellar tonsils herniation at the level of foramen magnum measured in sagittal, coronal and axial planes reflecting respectively the length, height and width of the herniated cerebellar tonsils using cranio-cervical MRI was measured.

Cerebellar tonsils were considered to be ellipsoid structures and its volume was calculated based on the following formula: volume =*lenght∗height∗widht∗*0.52(Adopted from radiological calculation of prostate volume and prostate ellipsoid formula)(17)(18). The most inlet of the foramen magnum which is the inner diameter of opisthion-basion plane was used as reference point for the above mentioned measurements. Foramen magnum was considered to be sphere shaped; the volume of foramen magnum at the largest point was calculated using the following sphere formula: *V*=4/3 *π r*3

## DATA MANAGEMENT AND STATISTICAL ANALYSIS

To assess the relationship between clinical manifestation of CMI and cerebellar tonsils, we computed various non-parametric statistical tests and hypothesis testing to analyze correlation and variation of cerebellar tonsils measurement in sagittal,coronal and axial planes, cerebellar tonsils volume, and the T/F volume ratio (cerebellar tonsils volume/foramen magnum volume) we run spearman rank correlation to analyze correlation between headache severity and both sagittal and coronal measurements (right and left), correlation between total cerebellar tonsils volume and foramen magnum, correlation between cerebellar tonsils volume and headache severity, myelopathy severity and tonsils volume, and T/F volume ratio and myelopathy. The hypothesis tests using Wilcoxon rank sum tests were also constructed to assess the significance of variation between right and left cerebellar tonsils measurements (coronal and sagittal measurements)in diagnosis of CMI, cerebellar volume (left and right), and sagittal and coronal measurements. we computed descriptive analysis to determine demographic characteristics of the study participants, occurrence of signs and symptoms among patients with CMI. Before running every test, we assessed the skewness of the data by variable and made a log-transformation whenever suitable likely, we found none. When we found that the data were skewed, we went on running and look for a suitable non-parametric tests. We did all the calculations in python 3 using scipy. stats, Wilcoxon, pearsonr, seaborn, and matplotlib.pyplot packages and pandas library

## RESULTS

Our study included ninth six (96) patients based on calculated sample size and statistical power of the study. the study results are presented on basis of the study objectives and statistical findings. CMHI was most common in female patients. Fifty-nine patients in ninety-six patients(59/96) representing 61.4% was female while the male patients with CMI was thirty-seven in ninth six patient(37/96) representing 38.5% of the study population. The majority of patients with CMI in our study was in fourth and fifth decade with age ranges : less than 20 years of age, between 20-30 years of age, between 40-50 years of age and Above 50 years representing 7.3%, 12.5%, 39.6%, 32.3% and 8.3 % respectively. The table1 below summarizes the results on demographic information of our study population.

Referring to the figure2 above, summarizing the finding on distribution of signs among CMI patients. Myelopathy followed by sensory loss were the common finding. Upper motor neuron lesion like Hoffman sign, spasticity and clonus was found to be 8% and 18% respectively. The motor weakness as result of CMI was found in 15% of our study population. Scoliosis as result of CMI was found in 5% of our study population. The Syringomyelia was defined as cerebral spinal fluid cavitation within the spinal cord seen as high signal intensity on T2W MRI image sequence and was found in 8% of our study population.

**Figure 1:**
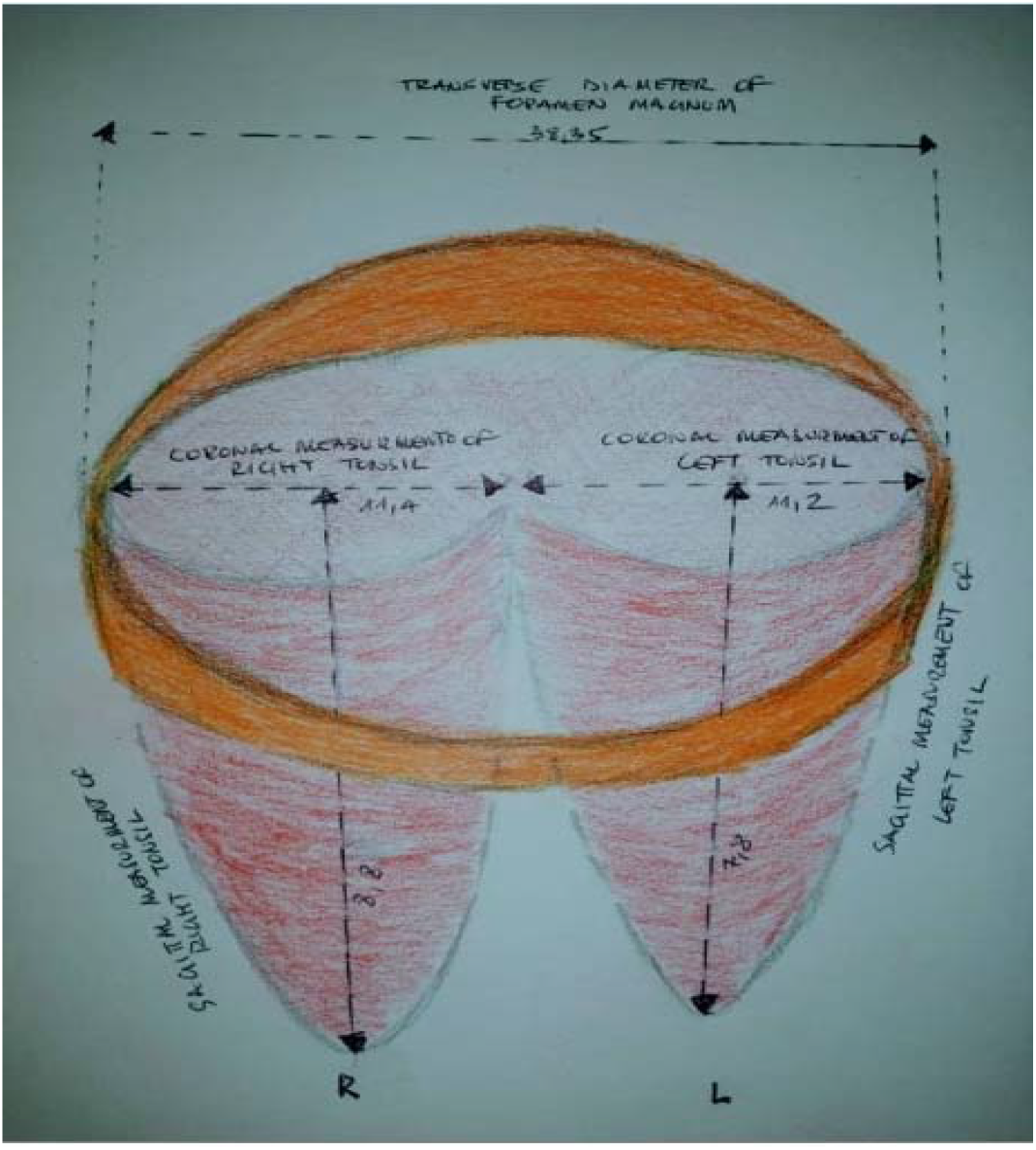
Illustrative drawing showing measurement of cerebellar tonsils in sagittal, coronal, axial plane and transverse diameter of foramen magnum used to calculate tonsil volume, foramen magnum volume and T/F volume ratio

**Figure 2:**
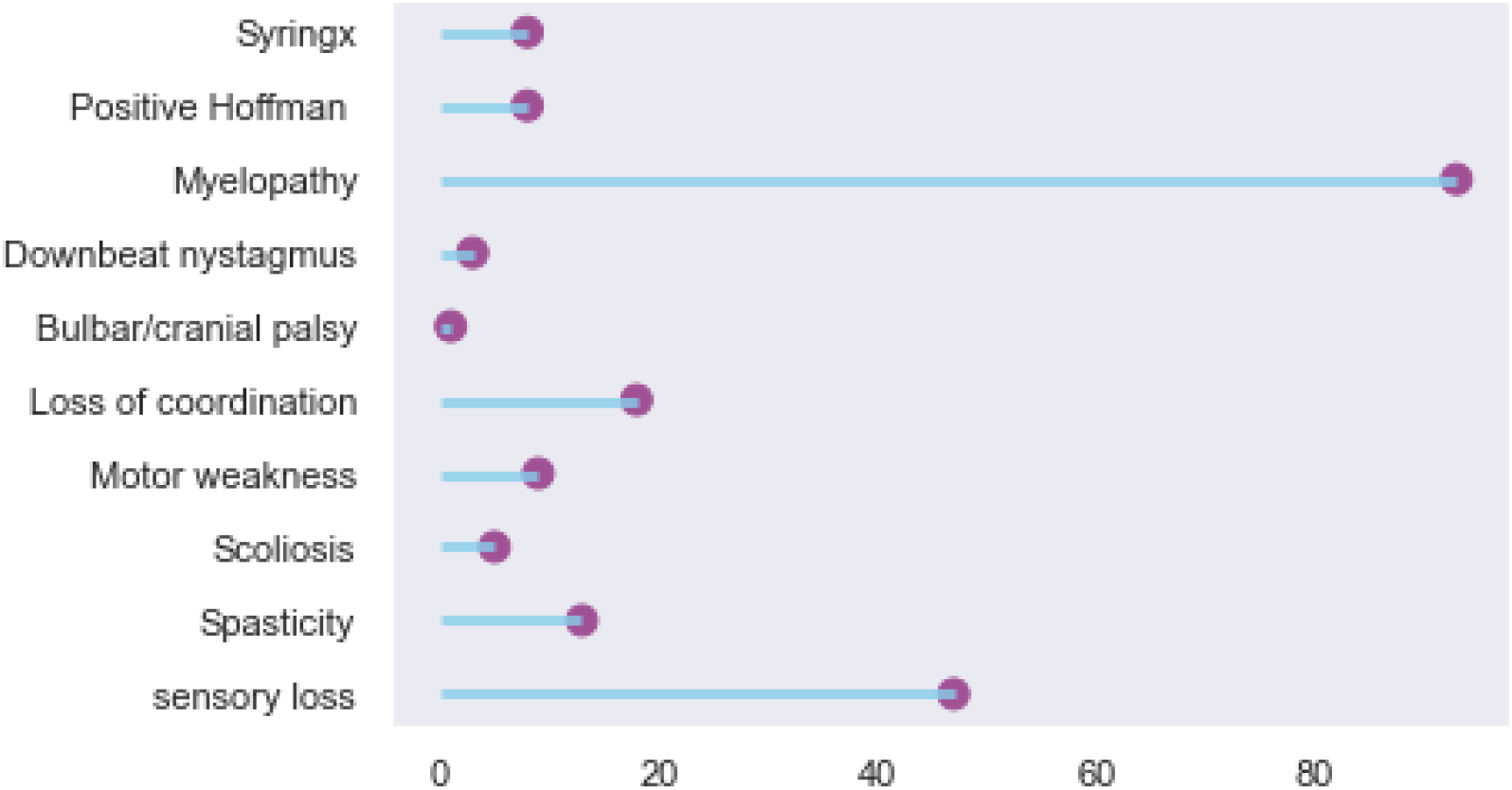
Distribution of signs among patients Chiari malformation type I.

Referring to the figure 3,4 showing headache and myelopathy severity according to NRS and modified Japanese Orthopedics Association (mJOA) respectively; patients with CMI mostly presented with severe headache, moderate headache with 58.3% and 41.7 % respectively according to pain numeric rating scale in our study. The severity of myelopathy is classified into mild, moderate and severe according to mJOA. Most patients with CMI was mild myelopathic with 81.2 % followed by moderate myelopathy with 11.5 % and severe myelopathy in 7.3% of study subjects.

**Figure 3:**
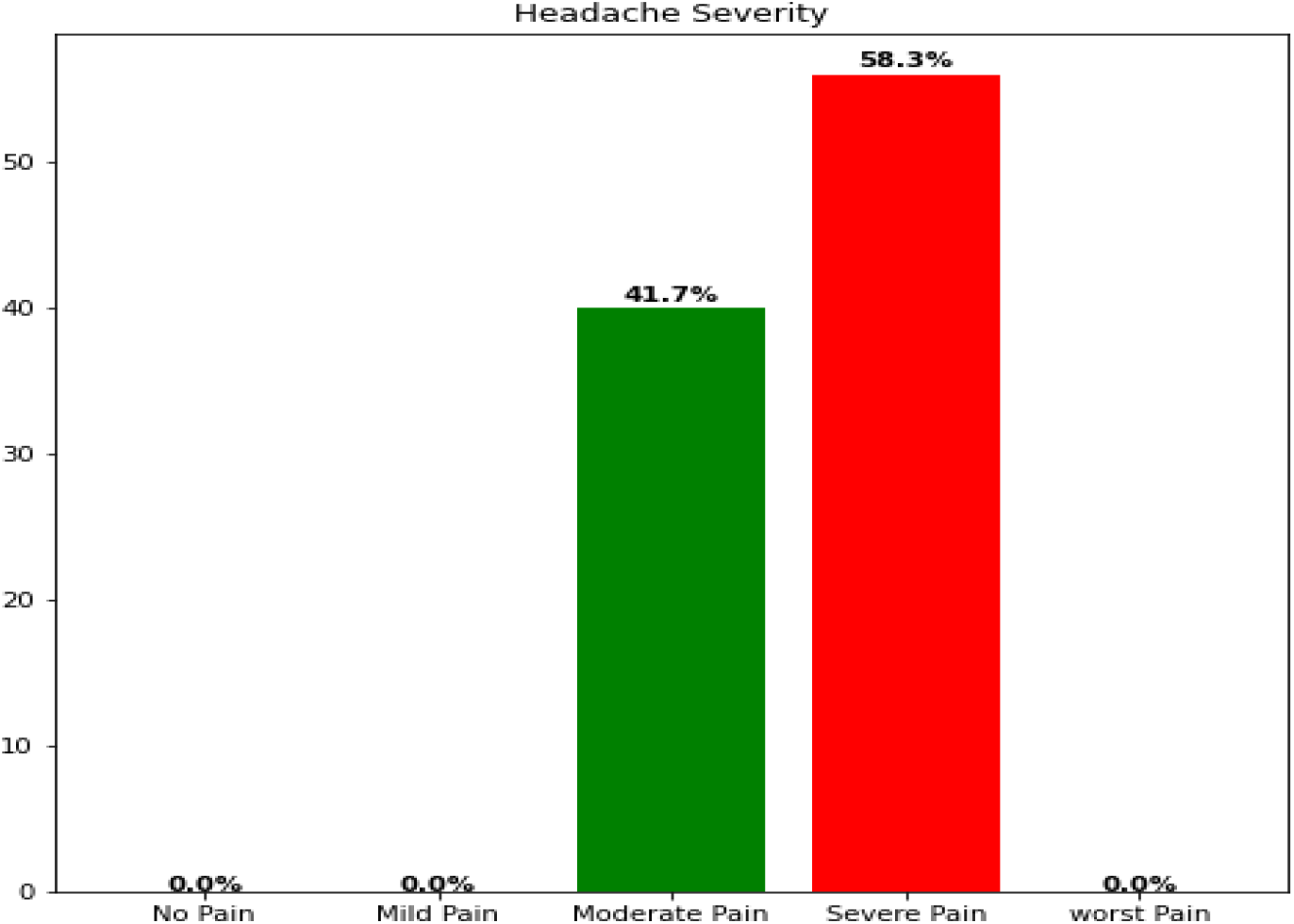
Headache severity according to pain Numeric rating scale(NRS) among CMI patients.

**Figure 4:**
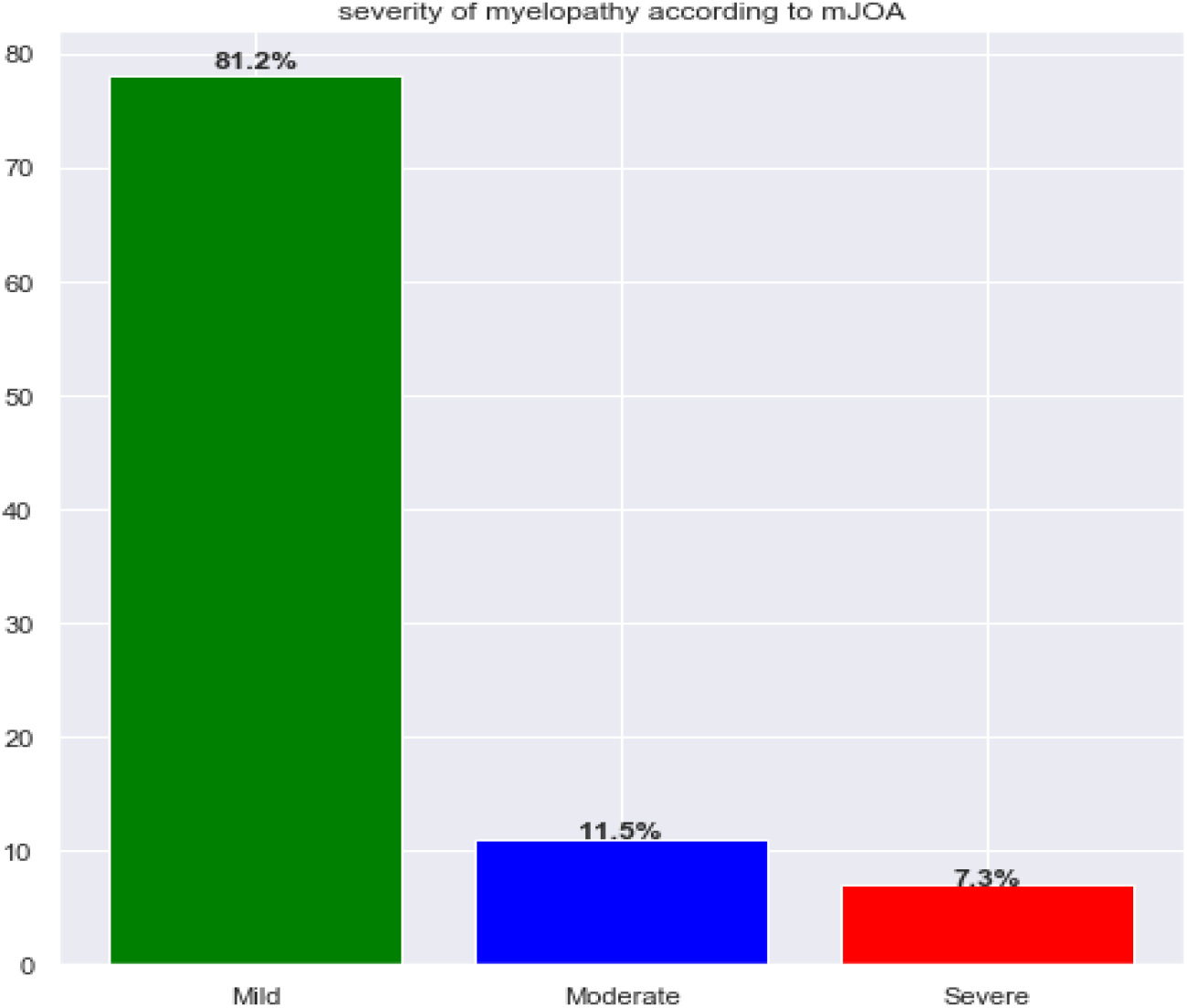
Myelopathy severity according to Modified Japanese Orthopedics Association (mJOA)

**Figure 5:**
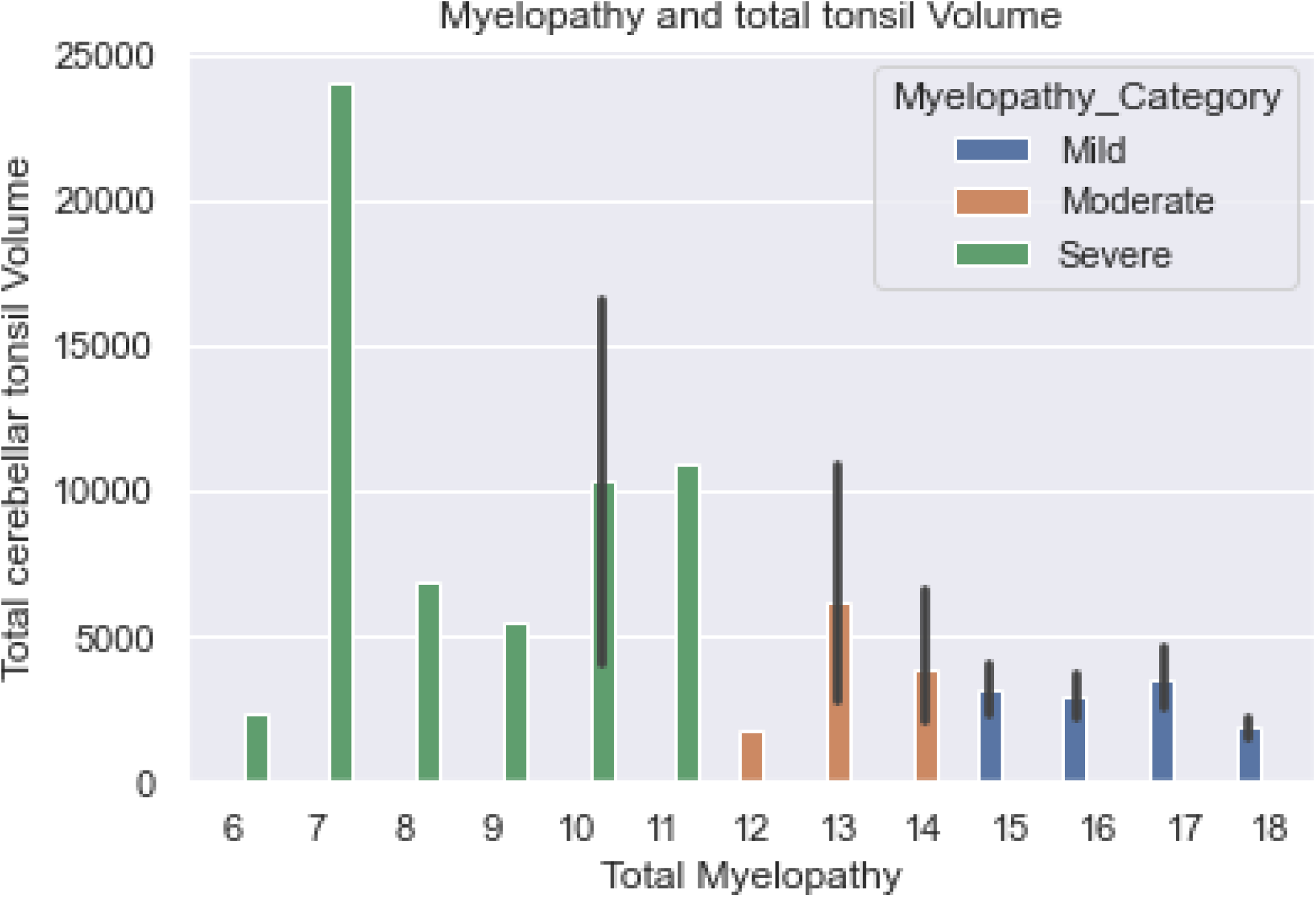
Correlation between myelopathy severity and cerebellar tonsils volume.

Referring to the table 2 summarizing tri-dimensional radiological measurements of cerebellar tonsils herniation and foramen magnum; the measurements of herniated cerebellar tonsils within foramen magnum was done in three dimensions; coronal measurement, sagittal measurement, axial measurement and volume of herniated cerebellar tonsils was calculated.

**TABLE 1:** Demographic information in patients with CMI.

|  | **Frequencies** | **Percentage (%)** |
| --- | --- | --- |
| **Sex** |  |  |
| Male | 37 | 38.5 |
| Female | 59 | 61.4 |
| **Age** |  |  |
| < 20 years | 7 | 7.3 |
| 20-30 years | 12 | 12.5 |
| 30-40 years | 38 | 39.6 |
| 40-50 years | 31 | 32.3 |
| Above 50 years | 8 | 8.3 |

**Table 2.** RADIOLOGICAL MEASUREMENT OF FORAMEN MAGNUM AND CEREBELLAR TONSILS.

|  |  |  |  |
| --- | --- | --- | --- |
| Coronal (mm) | Median(IQR) | Sagittal (mm) | Median(IQR) |
| left | 11.2(9.3 13.1) | left | 7.8(6.2 10.4) |
| right | 11.4(10.2 14.1) | Right | 8.8(7.2 11.3) |
| **Transverse Diameter of Forman magnum** |  |  |  |
| **Median (IQR)mm** |  |  |  |
| 38.35(36.2 40.63) |  |  |  |
| **Foramen Magnum volume** |  |  |  |
| **Median (IQR)mm<sup>3</sup>** |  |  |  |
| 29,517.3(24,774.6 35,088.1)mm<sup>3</sup> |  |  |  |
| **Cerebellar tonsils volume** |  |  |  |
| **Median (IQR)** |  |  |  |
| 2,427.61(1,706.01 3,625.2)mm<sup>3</sup> |  |  |  |
| **T/F volume ratio (Cerebellar tonsils volume/FM_ volume)** |  |  |  |
| **Median (IQR)** |  |  |  |
| 0.082(0.053 0.177) |  |  |  |

The diameter of FM was measured and the volume of FM calculated using circle formula and transverse diameter of FM diameter respectively.

The left median coronal measurement were11.2 mm with interquatile(IQR) range (9.3 mm-13.1 mm)

The right median coronal measurement were 11.4 mm with interquatile range of (10.3 mm-14.1mm)

The left median sagittal measurement were 7.8 mm with interquatile range of (6.2 mm - 10.4mm)

The right median sagittal measurement were 8.8 mm with interquatile range of (7.2mm-11.3mm) The median transverse diameter of FM were 38.25 mm with interquatile range of (36.2mm-40.63mm)

The median volume of foramen magnum were 29,571.3 mm^3^ with interquartile range of(24,774.6mm-35,088.1mm)

The median of total cerebellar tonsils volume were 2,427.61mm^3^ with interquartile range of (1,706.01mm^3^ - 3,625.2mm^3^)

The median and interquartile range of T/F volume ratio(Cerebellar tonsils volume/FM volume) were 0.082(0.053 0.177) respectively

Referring to the figure10 above showing the relationship between myelopathy severity and the volume of herniated cerebellar tonsils as well as the relationship between T/F volume ratio and myelopathy severity according to mJOA; there was a correlation between myelopathy severity and the volume of herniated cerebellar tonsils. The more patient has increased volume of herniated cerebellar tonsils, the more the severity of myelopathy on mJOA scale.

Referring to the figure 11 above; there is a crescendo relationship between T/F volume ratio (cerebellar tonsils volume/Foramen magnum volume) and severity of myelopathy. The increase of volume ratio is proportional to the severity of myelopathy on mJOA scale.

## DISCUSSIONS

The majority of patient with CMI in our study were in fourth and fifth decade with age ranges: less than 20 years of age, between 20-30 years of age, between 40-50 years of age and Above 50 years representing 7.3%, 12.5%, 39.6%, 32.3% and 8.3 % respectively. Our finding are similar to the finding of Aska Arnautovic and colleagues in their review of study on demographics, operative treatment and outcome showed that peak age of presentation in the pediatric studies were 8 years, followed by 9 years, and in the adult series,41 years, followed by 46 years old with female to male ratio 1.3:1(23) James R Houston et al in their study on evidence for sex difference in morphological abnormality in CMI report no sex difference of morphometric abnormalities of CMI malformation.(24)

Patients with CMI mostly presented with severe headache and moderate headache with 58.3% and 41.7 % respectively according to pain numeric rating scale in our study. There was no correlation between headache severity and cerebellar descent measurement in both sagittal and coronal. In contrast there was proportional correlation between headache severity and T/F volume ratio (cerebellar tonsils volume/foramen magnum volume ratio). Our finding are similar to the study by Dan S. Heffez et al who report no correlation between symptoms severity with extent of cerebellar descent in CMI patients (5).

Jonathan Pindrik et al in their study on clinical presentation of CMI and syringomyelia in pediatric population report headache or neck pain in 27%–70%, motor or sensory deficits of the extremities, 6%–17%,Scoliosis 18%–50%,Ataxia or gait impairment, decreased coordination 4%–9%,Spasticity 6% (27)

The syringomyelia is define as cerebral spinal fluid cavitation within the spinal cord seen as high signal intensity on T2W image MRI sequence. Syringomyelia was found in 8% of our study population.

The pathogenesis of syringx associated with CMI has not been clarified. Many theories explaining the presence of syringx in CMI have been postulated in literature, however disorder in CSF pathway around FM is taught to be main cause of syringx (28)

The prevalence of syringx in CMI is variable among adults and pediatric population. The occurrence of a syringx in association with CMI varies between 35 to 75% in pediatric patients while in adults syringx associated with CMI varies between 40 to 60% (26).

Aitken and colleagues identified spinal syringomyelia in 12% of patients with CMI(29). The scoliosis associated with CMI in our study is 5% of the study population.

The exact mechanism of scoliosis in association with CMI is not well known. It is thought to result from chronic compression of cervico-medullary junction by cerebellar tonsils, abnormality in posterior column tracts affect postural reflex resulting in difficulty sustaining appropriate posture their by subsequent scoliosis. However others authors associated the scoliosis with the presence of syringomyelia(30)

Our finding are similar to the result of Mohammad Hassan A.et al who report scoliosis resulting from isolated CMI in 3.0% (30). On the other hand Palma Ciaramitaro et al found scoliosis in 18%–50% of CMI(26)

Siri Sahib S. et al in their study on morphometric and volumetric in pediatric population with CMI found no clinical usefulness of 2D or 3D measurements that can help to differentiate symptomatic pediatric patients with CM-I from those who are asymptomatic CMI in their study hence, recommend an other diagnostic criteria beyond classic measurement of tonsil position useful for detecting best candidates for surgical management in patients with CMI(33). Our study showed a correlation between myelopathy severity and the volume of herniated cerebellar tonsils. There was a proportional increase of volume of herniated cerebellar tonsils and the severity of myelopathy. In addition, our study found a crescendo relationship between T/F volume ratio (cerebellar tonsils volume/foramen magnum volume) and severity of myelopathy. The increase of T/F volume ratio is proportional to the severity of myelopathy.

Currently, there is paucity of data in literature showing the relationship of severity of myelopathy and cerebellar tonsil volume as well as the relationship between T/F volume ratio (cerebellar tonsils volume/foramen magnum volume) and severity of myelopathy as discussed in our study.The T/F volume ratio (cerebellar tonsil volume/Foramen magnum volume) and cerebella tonsil volume can be an additional tool to be used in diagnosis and predict the severity of CMI.

## CONCLUSIONS

Many studies have showed no correlation between cerebellar tonsils herniation and clinical severity in patients with CMI and suggests other diagnostic criteria beyond standard measurement of cerebellar tonsil position which can help in determining the best candidates who need surgical management for patients suffering from CMI. Our study is the first in literature to show the correlation between the tonsil volume and T/F volume ratio(cerebellar tonsil volume/foramen magnum volume) and headache severity using pain numeric rating scale(NRS) as well as severity of myelopathy.

Our study shed light on cerebellar tonsil volume and T/F volume ratio as predictors of severity in patients with CMI.

Further study is recommended to find out if cerebellar tonsils volume and T/F volume ratio(cerebellar tonsil volume/foramen magnum volume) can be used as diagnostic tool in patients with CMI.

It is of great importance to calculate both sagittal and coronal measures in assessment of patients with CMI.

## Data Availability

All data produced in the present study are available upon request to the authors

## LIST OF ABBREVATIONS

CMI: Chiari Malformation type I
CSF: Cerebral Spinal Fluid
MRI: Magnetic Resonance Imaging
CT: Computed tomography
VAS: Visual Analog Scale
NRS: Numeric rating scale
m JOA: Modified Japanese Orthopedic Association
NRS: Numeric rating scale
CHUK: Centre Hospitalier Universitaire de Kigali.
KFH: King Faisal Hospital
RMH: Rwanda Military Hospital
UR: University of Rwanda
CMHS: College of Medicine and Health Sciences.
IRB: Institutional Review Board
MC: Rae line
FM: Foramen magnum
CSI: Chiari Severity index
T/F volume ratio: Cerebellar tonsils volume /Foramen magnum volume

